# Patient Experience with Amputation or *Limb Loss*: Literature Review

**DOI:** 10.1101/2022.04.23.22274207

**Authors:** Atyanti Isworo, Moses Glorino Rumambo Pandin

## Abstract

**Backgrounds:** Amputation which is the removal of body parts, the reality faced by patients who must be amputated raises various responses ranging from rejection to acceptance. The process of each patient is certainly different according to the experience experienced by each patient.

**Methods:** This literature review uses articles published from 2018-2022 with source from databased ScienceDirect, Pubmed, Scopus, Sage Journals, EBSCO-Host, BMC Journal, Springer Link, Taylor and Francis, and ProQuest. The keywords used are patient experience, perspectives of patients, amputations, limb loss, low extremity amputation, qualitative, qualitative study. Article selection follows “PRISMA” flow.

**Results:** This systematic review have a total of 6 articles meet the criteria for analysis. Respondents involved in this qualitative research amounted to 109 respondents who had undergone the amputation process. There are several themes that are used to explore the patient’s experience, the themes used are divided into several aspects in the form of aspects of physical changes, aspects of psychological changes, and also aspects of social adaptation that patients must go through after undergoing the amputation process.

**Conclusion:** Through a qualitative study, it was found that the average patient expressed feelings of worry, fear, anxiety, feeling useless, no longer liking his body, feeling lost in self-confidence, and no longer wanting to interact socially because of the negative response from the surrounding environment towards the patients.

## INTRODUCTION

Amputation is a surgical procedure that has been quite popular for a long time, the development of surgical techniques and prosthetic design began as a result of war. The term amputation comes from the Latin “amputare” which means cutting. Until now amputation was defined as the removal of a limb as a result of trauma (traumatic amputation) or in an attempt to control disease or disability (therapeutic amputation) (Daniel & Nicolle 2012). Amputation, which is the removal of a body part, is often done on the extremity. Lower extremity amputation is often required because of progressive peripheral vascular disease (diabetes type 2), fulminant gas gangrene, trauma (crushing injury, burns, frostbite, electric burns, blasts, and ballistic injuries), congenital defects, chronic osteomyelitis, or malignant tumors. Of all these disease states, peripheral vascular disease accounts for the majority of lower extremity amputations. Upper extremity amputation is less common than lower extremity and is most often required because it is good for treating traumatic wounds and malignant tumors (Smeltzer & Bare 2013). The amputation rate in the world is 0.7 per 1000 population, while in Asia is 31 per 1000 population. Based on data, the incidence of amputations in Indonesia in 2010-2011 increased from 35.5% to 54.8%. (Purwanti 2014).

Patients who have undergone the amputation process will experience several condition responses. Kubler-Ross divides these responses into several stages such as rejection, anger, bargaining, depression, and self-acceptance. Therefore, the patient needs to go through the depression stage and reach the final stage, which is self-acceptance (Lind et al. 2014). Self-acceptance refers to one’s life satisfaction and happiness which is very important for good mental health. Someone who is able to accept themselves understands their strengths and weaknesses. Effective self-acceptance consists of 1) having an accurate perception of reality, 2) being able to cope with or dealing with stress and anxiety, 3) having a positive self-image, 4) being able to express feelings, and 5) having relationships good interpersonal (Hasan, Lilik & Agustin 2013).

The grieving process that must be passed will allow a person to accept the situation and live life, and self-acceptance can improve the quality of life for amputees. However, besides that, there is a range of responses experienced by amputees, some experience frustration and death anxiety, some of which stop at the depression stage. Amputees who stop at the depression stage actually worsen the patient’s condition, learned helplessness will occur so that the amputee does not have the desire to live (Lind et al, 2014). This adaptation process is a different process for each patient who undergoes amputation. With the diversity of responses and experiences that are certainly different for each person, researchers are interested in reviewing several previous studies that have explained the results of responses or experiences of patients undergoing the amputation process with qualitative studies that explained in detail the experiences and perceptions of amputees about their condition.

## METHOD

### Eligibility Criteria

Determination of the article criteria used the PICOS framework search strategy (Population, Intervention, Comparison, Outcome, Study type) according to the inclusion criteria and exclusion criteria that have been determined as in the table below.

| Criteria | Inclusion | Exclusion |
| --- | --- | --- |
| *Population* | Adult | Children and elderly |
| *Intervention* | Nursing intervention | No exclusion criteria |
| *Comparison* | No comparison | No exclusion criteria |
| *Outcome* | Experiences of patients who undergo amputation or limb loss | Does not discuss the experiences of patients who undergo amputation or limb loss |
| *Study type* | Qualitative study | Cross sectional study, mixed method, quasi experimental design, clinical trial or randomized controlled trial, Systematic or literature reviews |

### Search Results

The search begins by specifying keywords using conjunctions (AND, OR NOT or AND NOT). *Keywords used*: (“*patient experience”* OR “*perspectives of patients*”) AND (“*amputations”* OR “l*imb loss”* OR “*low extremity amputation”*) AND (*“qualitative”* OR *“qualitative study”*). Initial search results started by searching the ScienceDirect database, obtaining 593 articles, Scopus obtaining 34 articles, Sage Journals obtaining 249 articles, EBSCO-Host obtaining 232 articles, BMC Journal obtaining 360 articles, Taylor and Francis obtaining 241 articles, PubMed obtaining 115 articles, *Springer Link* obtaining 160 articles, and *ProQuest obtaining* 252 articles. The total titles obtained from this search amounted to 2236 articles, which were then entered into the Web Importer (Mendeley) to eliminate duplicate articles. After that, the screening stage was carried out on the titles and abstracts of articles that did not match the research topic. After going through the screening stage, then an assessment of the feasibility of the article is carried out based on the completeness of the complete downloadable article. After that is the screening stage based on the inclusion and exclusion criteria that have been set in the study so that 6 articles are found that are deemed to meet the criteria for further analysis according to the established theme. A summary of the articles that have been analyzed is listed in the PRISMA (Preferred Reporting Item for Systematic reviews and Meta-Analyses) diagram below:

**Figure 1.**
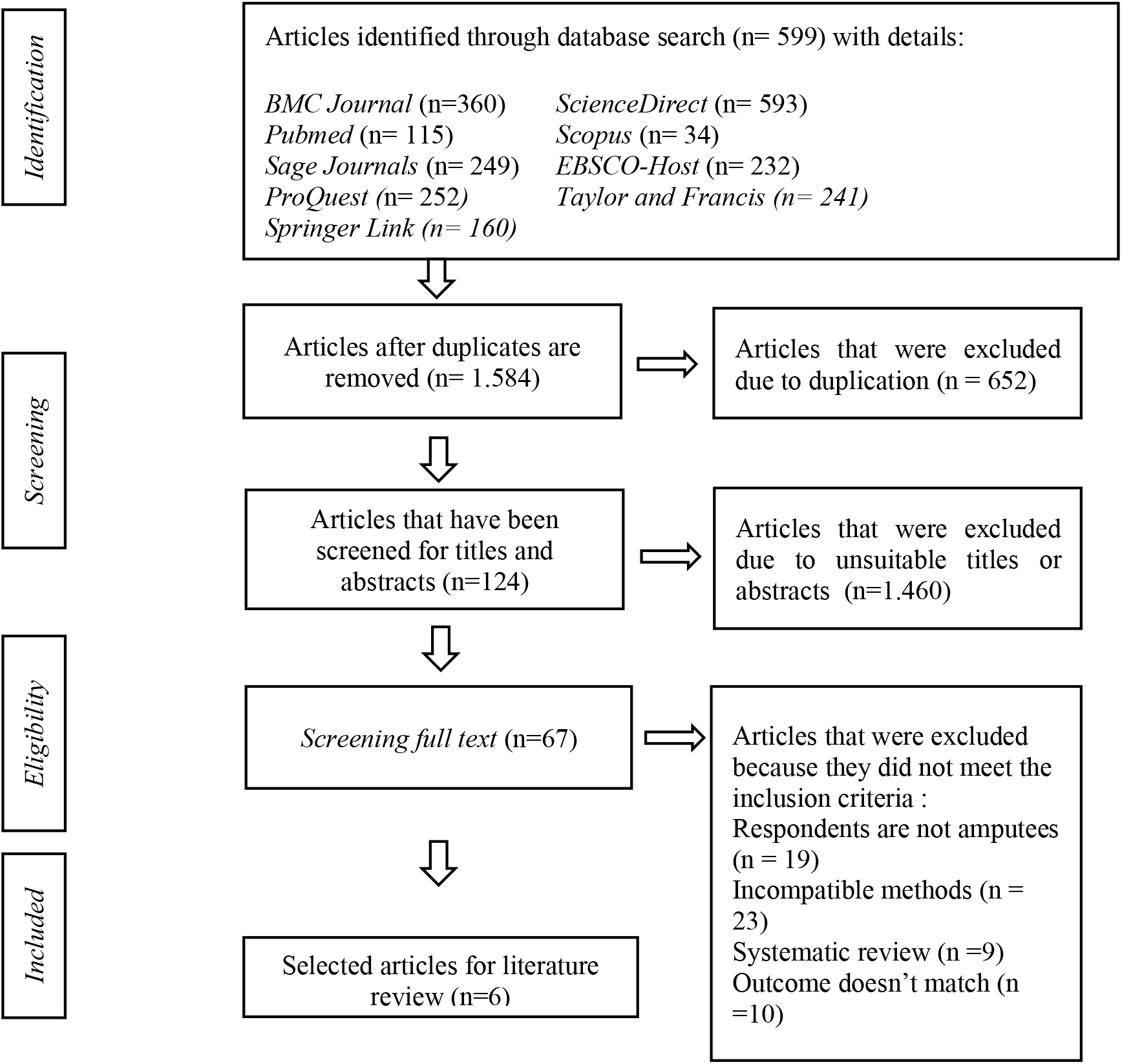
Flowchart PRISMA.

## RESULT

**Table 2.** Literature search result.

| No | Title, Author, Year | Method | Result |
| --- | --- | --- | --- |
| 1. | Experiences and needs of patients with lower limb amputation in Saudi Arabia: a qualitative study (Abouammoh, Aldebeya & Abuzaid 2021) | Design: qualitative<br>Sample: 13 patients<br>Variable: patients' experience<br>Instrument: semi structured interview questionnaire sheet<br>Analysis: 3 themes | Hopelessness and depression, impaired body image, religious attitudes, and family and community support all contribute to shaping the overall patient experience, including psychological and physical adjustment. |
| 2. | Early Patient Experiences of Primary Above-the-Knee Amputation From Vascular Etiologies: A Phenomenological Study (Gómez-ibáñez et al. 2021) | Design: qualitative<br>Sample: 20 patients<br>Variable: patients' experience<br>Instrument: semi structured interview questionnaire sheet<br>Analysis: 3 main categories | Disclosing based on 3 main categories, fear, available health resources, and also the influence on family and social life. The fear and anxiety felt by patients due to amputation will depend on several factors including physical, psychological, and social aspects. |
| 3. | Patient Experience of Recovery After Major Leg Amputation for Arterial Disease (Columbo et al. 2018) | Design: qualitative<br>Sample: 20 patients<br>Variable: patients' experience<br>Instrument: structured interview guide<br>Analysis: 2 themes | Describe unmet needs and commonly used coping strategies. Participants felt under-treated included uncontrollable pain, feeling unprepared to live with the amputation, and questions about prosthetics. Two of the 5 focus group participants expressed a preference for amputation earlier in the treatment course. |
| 4. | "I Wish I Could Have My Leg ": A Qualitative Study on the Experiences of Individuals With Lower Limb Amputation (Seyman & Ozcetin 2022) | Design: qualitative<br>Sample: 12 patients<br>Variable: patients' experience<br>Instrument: semi structured interview questionnaire<br>Analysis: 3 themes | The main three themes described in the research are loss of control over one's own life, dreams versus reality, and perception of the future. Most of the participants emphasized that they were facing unwanted experiences in their post-amputation life. |
| 5. | Struggling for normality : experiences of patients with diabetic lower extremity amputations and post-amputation wounds in primary care (Zhu et al. 2022) | Design: qualitative<br>Sample: 9 patients<br>Variable: patients' experience<br>Instrument: Verbatim, in-depth interview<br>Analysis: IPA (interpretative phenomenological analysis) | The essential meaning of the phenomenon 'life experiences of diabetic patients with lower extremity amputation and post-amputation injuries' can be interpreted as a choice to strive for a normal life which includes four common sense domains: physical loss of normality is disturbed, emotional impact worsens disturbed normality, further social challenges provoke normality disturbed, and attempts to get back to normal. |
| 6. | A qualitative study exploring individuals' experiences living with dysvascular lower limb amputation (Mackay et al. 2020) | Design: qualitative<br>Sample: 35 patients<br>Variable: patients' experience<br>Instrument: semi structured interview questionnaire sheet, audio recording, and translated by verbatim<br>Analysis: Thematic | Thirty-five interviews were completed with individuals with dysvascular lower extremity amputations. Participants described lower extremity amputations as having an impact on many aspects of their lives, resulting in changes in mobility, social activities and roles, and psychological well-being. The three main factors that shape an individual's experience with dysvascular lower extremity amputations include social support, accessibility, and socioeconomic factors. |

## DISCUSSION

Based on the results of the discussion of several studies above, the experience of patients undergoing amputation shows that efforts are needed for physical and psychosocial adjustments, faster wound healing, higher coping efficacy, and higher levels of social adaptability to return to normal (Zhu et al. 2022). On average, patients experience pressure from both internal and external factors, including anxiety, hopelessness, depression, body image disturbances, loss of confidence in their abilities, lack of family support and also lack of support from their social environment (Abouammoh, Aldebeya & Abuzaid 2021).

Moreover, there are several emotional responses from the experience of patients undergoing amputation, which are divided into three main categories: 1) Fear, where the patient expresses fear of not being able to walk again after the amputation, the fear of always being dependent on others and not being able to meet their own needs, and the intensity of pain and phantom limb syndrome that will be felt. “How can I experience itchy and sore if I don’t have toes or feet anymore? Yes, I noticed the heels. I noticed how my socks tighten even I don’t have feet. Don’t tell me it’s funny. But one thing is clear, what I feel is not my feet, I mean how it used to be, I don’t know what I feel, but it’s not my feet.” (P1) (Gómez-ibáñez et al. 2021). 2) Concerns, the patient expressed concern about the recovery process that he will undergo, adaptation while at home, adaptation to the use of a wheelchair, and even adaptation to the use of prosthetic limbs which may require a long process. “Three or four months after, the absolute worst… I literally sat in the kitchen crying over how my life had changed so much. ‘What will I do to support my family, and how will I take out the trash?” (P2) (Columbo et al. 2018). 3) Feeling hopeless and useless that leads to depression, the patient expresses his feelings about how the attitude of the people around him after undergoing the amputation, the patient expressed a lot of people look down on him and don’t even give him support, hence the patient is aloof and doesn’t do social activities. “I don’t know how I can go back to work and meet my young students… I don’t want to hear negative comments…” (P3) (Abouammoh, Aldebeya & Abuzaid 2021). “Everything has changed. I used to be able to wake up from bed, but now I can’t. This might be simple for a lot of people, but it’s important for me. I can’t be someone who I used to be” (P4) (Seyman & Ozcetin 2022). “I think one of the biggest things I’m facing is social isolation. I’m no longer really out in the community, except for doctor’s appointments and grocery shopping. I used to be very active, I used to volunteer in many different places, I used to walk a lot every day” (P5) (Mackay et al. 2020).

Amputation is a choice of action that is not easy, this choice is clearly made based on serious and comprehensive considerations for both internal and external aspects of the patient. It can be said that the depressive reaction is a normal reaction to amputation rather than ordinary depression. However, a study by Abouammoh, Aldebeya & Abuzaid (2021) demonstrated that depressive reactions can be minimized by patient education as well as meeting with other amputees. This is the most useful source of information and support for patients and is considered more reliable by patients so that patients are better able to manage their depression by sharing their experiences.

According to other studies, there are several aspects that become themes to explore the feelings and experiences of patients who undergo amputation, which are divided into 4 themes in the form of physical loss that interferes with normality, bad emotional impact due to disturbed normality, higher social challenges after disrupted normality, and the attempt to regain normality after amputation (Zhu et al. 2022).

This unexpected change forces patients who have undergone the amputation process to face the fact that life can no longer be the same as before. As the participants pointed out, before the amputation, they focused on the pain they were experiencing and their chances of survival. However, they are forced to face the reality and difficulty of the recovery process after amputation. Furthermore, the participants’ comments described the important role the body plays in their daily experiences. Although most of the participants felt the need to move on with their lives, they also reported adaptation problems with experienced loss of physical function (Seyman & Ozcetin 2022).

For most patients, family members are their main companions who travel with them in their recovery process. After leaving the hospital to go home, one of the participants expressed feelings of anxiety, disorientation, and did not even acknowledging his own home (Zhu et al. 2022). The role of the family that helps and supports the patient’s adjustment after amputation is considered quite helpful and reduces the patient’s anxiety about the changes in the family he is facing (Mackay et al. 2020).

Overall, the results showed that the participants had similar challenges in adapting to their new life, although they believed that it was very important for them to learn to cope with their situation. These challenges had an important impact on their perception of amputation. In addition, participants felt that they had lost control of their lives, experienced shock over their situations, and had negative perceptions of the future (Seyman & Ozcetin 2022; Columbo et al. 2018).

## CONCLUSION

Amputation is a medical treatment option that requires serious consideration and affects all aspects of the patient. Patients who undergo amputation will indirectly experience changes afterward. In research that explores the experiences of patients after undergoing amputation in depth through qualitative studies, it was found that the average patient expressed feelings of worry, fear, anxiety, feeling useless, no longer liking their physical body, feeling lost in self-confidence, and no longer wanting to interact socially because response of the surrounding environment that tends to be bad towards it. Sufferers also complain that the influence of the family is very supportive and helps them to recover during the recovery process rather than having to struggle alone. This uneasy process of change makes some sufferers still try to build self-confidence and self-esteem to survive after the amputation process they go through.

## Data Availability

All data produced in the present work are contained in the manuscript

## LIMITATION AND RECOMMENDATION

A limitation of this review is the limited search of the literature for qualitative studies that discuss the experiences of patients undergoing amputation. This literature review is expected to be used as a guide in providing nursing interventions to patients who undergo amputation in the form of health education, empowerment, and support.

## ACKNOWLEDGEMENT

The authors would like to thank all the researchers whose articles were used for the discussion in this paper.

## REFERENCE

Abouammoh, N., Aldebeya, W. & Abuzaid, R. 2021, ‘Experiences and needs of patients with lower limb amputation in Saudi Arabia: a qualitative study’, Eastern Mediterranean health journal = La revue de sante de la Mediterranee orientale = al-Majallah al-sihhiyah li-sharq al-mutawassit, vol. 27, no. 4, pp. 407–13.

Columbo, J.A., Davies, L., Kang, R., Barnes, J.A., Leinweber, K.A., Suckow, B.D., Goodney, P.P. & Stone, D.H. 2018, ‘Patient Experience of Recovery After Major Leg Amputation for Arterial Disease’, Vascular and Endovascular Surgery Journal, vol. 52, no. 4, pp. 262–8.

Daniel, R. & Nicolle, L.H. 2012, Contemporary Medical Surgical Nursing (2nd ed.), Cengage Learning, Newyork.

Gómez-ibáñez, R., Granel, N., Watson, C.E. & Escribano, X. 2021, ‘Early Patient Experiences of Primary Above-the-Knee Amputation From Vascular Etiologies : A Phenomenological Study’, Clinical Nursing Research, vol. 30, no. 5, pp. 539–47.

Hasan, A., Lilik, S. & Agustin, R.W. 2013, ‘Hubungan Antara Penerimaan Diri dan Dukungan Emosi dengan Optimisme pada Penderita Diabetes Mellitus Anggota Aktif PERSADIA (Persatuan Diabetes Indonesia) Cabang Surakarta’, Jurnal Ilmiah Psikologi Candrajiwa, vol. 2, no. 2, pp. 60–74.

Lind, M., Svensson, A.M., Kosiborod, M., Gudbjörnsdottir, S., Pivodic, A., Wedel, H. & Rosengren, A. 2014, ‘Glycemic control and excess mortality in type 1 diabetes’, New England Journal of MedicineEngland Journal of Medicine, vol. 371, no. 21, pp. 1972–82.

Mackay, C., Cimino, S.R., Guilcher, S.J.T., Mayo, A.L., Devlin, M., Dilkas, S., Payne, M.W., Viana, R., Sander, L., Devlin, M., Dilkas, S., Payne, M.W., Viana, R. & Hitzig, S.L. 2020, ‘A qualitative study exploring individuals ‘ experiences living with dysvascular lower limb amputation’, Disability and Rehabilitation, vol. 1, no. 1, pp. 1–9.

Purwanti 2014, ‘Hubungan Motivasi dengan Efikasi Diri Pasien DM Tipe 2 dalam Melakukan Perawatan Kaki Di Wilayah Kerja Puskesmas Ponorogo Utara.’, Jurnal Ilmu Kesehatan, vol. 11, no. 1, pp. 68–77.

Seyman, C.C. & Ozcetin, Y.S.U. 2022, ‘“ I Wish I Could Have My Leg “ : A Qualitative Study on the Experiences of Individuals With Lower Limb Amputation’, Clinical Nursing Research Journal, vol. 31, no. 3, pp. 509–18.

Smeltzer, S.C. & Bare, B.G. 2013, Keperawatan Medikal Bedah Brunner dan Suddart Volume 1, Edisi 8., EGC, Jakarta.

Zhu, X., Goh, L.J., Chew, E., Lee, M. & Bartlam, B. 2022, ‘Struggling for normality : experiences of patients with diabetic lower extremity amputations and post-amputation wounds in primary care’, Primary Health Care Research & Development, vol. 21, no. 63, pp. 1–10.

